# Bridging the Coverage Gap: State Medicaid Limitations for Cardiac Rehabilitation Programs and the Risk to Disadvantaged Communities

**DOI:** 10.64898/2026.04.03.26350136

**Authors:** J. Curran Henson, Garrett S. Spears, Brett K. Daughdrill, Joshua N. Hagood, Srikanth Vallurupalli

**Affiliations:** Department of Internal Medicine, University of Arkansas for Medical Sciences, Little Rock, AR; College of Medicine, University of Arkansas for Medical Sciences, Little Rock, AR; Division of Cardiovascular Medicine, Department of Internal Medicine, University of Arkansas for Medical Sciences, Little Rock, AR

**Author notes:** Corresponding Author: *Srikanth Vallurupalli, MD*, Division of Cardiovascular Medicine, Department of Internal Medicine, University of Arkansas for Medical Sciences, 4301 W. Markham St. Little Rock, AR 72205.

## Abstract

**Background:** Cardiac rehabilitation (CR) is a cost-effective, evidence-based intervention that improves outcomes for patients with heart failure (HF), yet access remains inequitable, particularly among Medicaid enrollees. We evaluated state-level variability in Medicaid coverage for CR services and examined the implications for health equity in vulnerable populations.

**Methods:** We conducted a cross-sectional policy analysis of all 50 U.S. states to assess Medicaid coverage for outpatient CR services billed under CPT codes 93797 (without ECG monitoring) and 93798 (with ECG monitoring). Publicly available Medicaid documents were reviewed and supplemented with direct communication with state Medicaid agencies. States were categorized into full, partial/inconclusive, or no coverage. Geographic trends were visualized through heat maps and contextualized using state-level Medicaid enrollment data.

**Results:** Marked disparities in CR coverage were identified. Only 41 states reimbursed for CPT 93797, and 43 for CPT 93798. Eight states lacked coverage for either code, predominantly in the South and Mountain West, including Arkansas, Georgia, Louisiana, Mississippi, Nevada, and Utah. States with the highest Medicaid enrollment (e.g., Louisiana, Arkansas) often provided no CR coverage, compounding access barriers for high-risk, low-income populations.

**Conclusions:** The absence of standardized Medicaid coverage for CR contributes to systemic inequities in cardiovascular care, disproportionately impacting disadvantaged communities. Aligning Medicaid policies to ensure universal CR access—particularly through tele-rehabilitation and value-based care models—could reduce hospitalizations, improve survival, and promote health equity across the U.S.

## Introduction

Heart Failure (HF) is a multifactorial disease that causes significant burden at many levels of the public health spectrum. According to the Heart Failure Society of America, HF is a growing public health concern [1], affecting over 6.7 million adults in the United States, with projections estimating this number could surpass 11.4 million by 2050. The projected increase in HF prevalence is expected to impose a substantial economic burden, with total annual costs exceeding $160 billion by 2030, including direct medical expenses and indirect costs such as lost productivity [2]. If all costs of cardiac care for heart failure patients are attributed to heart failure, the projected direct costs could be as high as $160 billion [2], making heart failure one of the most expensive medical conditions in contemporary society.

Among the most cost-effective, evidence-based strategies to mitigate this burden is outpatient cardiac rehabilitation (CR), particularly under CPT codes 93797 (without ECG monitoring) and 93798 (with ECG monitoring) [3]. CR provides documented reductions in all-cause mortality (by 25–30%) and hospital readmissions (by as much as 31%) [5,10]. Yet despite its benefits, utilization remains abysmally low among Medicaid enrollees, with under referral and non-completion rooted in socioeconomic and geographic barriers [8,11].

Since the Centers for Medicare & Medicaid Services (CMS) approved CR for patients with stable HF in 2014 [6], widespread uptake has been anticipated; however, Medicaid policies vary significantly by state, reflecting both administrative discretion and political priorities [7]. This patchwork coverage structure leaves many low-income individuals—especially those in non-expansion states—without access to lifesaving rehabilitation care [9]. The lack of Medicaid coverage for CR is not a neutral policy—it tends to systematically exclude the very populations that face the highest burden of disease, particularly in southern and midwestern states with both higher HF prevalence and lower Medicaid generosity [9,12]. This variation in coverage has disproportionate consequences for vulnerable populations, where Medicaid is often the primary or sole source of insurance for rural, minority, and low-income individuals, and these disparities extend beyond economic barriers. Notably, Black Americans are nearly twice as likely to develop HF before the age of 65 and are significantly less likely to complete CR when referred [11,13]. Coupled with lower rates of Medicaid expansion, this creates a compounded public health threat in states where social determinants of health, including socioeconomic status, access to healthcare, racial disparities, and limitations in Medicaid coverage converge [14,15]

## Methods

We conducted a comprehensive cross-sectional, 50-state policy analysis to evaluate outpatient cardiac rehabilitation (CR) coverage for patients insured solely through Medicaid. Specifically, we examined coverage for CR services under CPT code 93797 (without continuous ECG monitoring) and CPT code 93798 (with continuous ECG monitoring), which represent the standard billing codes for structured, outpatient CR programs.

Our data collection began with a detailed review of publicly available Medicaid fee schedules, provider manuals, and reimbursement documents hosted on each state’s official Medicaid website. This initial online research was supplemented with direct outreach to state Medicaid agencies to verify or clarify coverage status when public data were incomplete, outdated, or ambiguous. Phone interviews were conducted with Medicaid representatives from these states to confirm the accuracy of available documentation and obtain updated information as needed.

States were then categorized into three groups based on their CR coverage: (1) Full Coverage, states covering both CPT 93797 and 93798; (2) No Coverage, states with no evidence of Medicaid reimbursement for either code; and (3) Partial or Inconclusive, states where only one code was covered or where coverage status could not be confirmed through available documentation or agency contact.

To illustrate geographic trends and disparities, we developed U.S. heat maps representing state-level CR coverage, highlighting regions where Medicaid beneficiaries face reduced or absent access to evidence-based secondary prevention services. This approach allowed us to identify and compare policy gaps across states, particularly in regions with high cardiovascular disease burdens and socioeconomically vulnerable populations.

## Results

Our review of Medicaid CR coverage across all 50 U.S. states revealed significant variation in outpatient cardiac rehabilitation reimbursement, with notable geographic disparities that align with broader patterns of health inequity. Specifically, we found that CPT code 93797, which refers to supervised exercise therapy without continuous electrocardiographic (ECG) monitoring, was reimbursed in full by 41 states. However, eight states: Arkansas, Georgia, Iowa, Louisiana, Mississippi, Nevada, New Jersey, and Utah offered no coverage at all for this code (Figure 1). In addition, one state, Kansas, presented incomplete or ambiguous information, leading us to classify this state’s status as partial or inconclusive. Two states, Tennessee and Kansas comprised systems that partnered with private insurers which offered a group of subsidized plans through each state’s respective Medicaid agency. Tennessee’s Medicaid disbursement through public-private partnership with 4 private insurers were able to be verified for CR coverage, but Kansas data was unable to be accessed through all of the private insurer data platforms.

**Figure 1:**
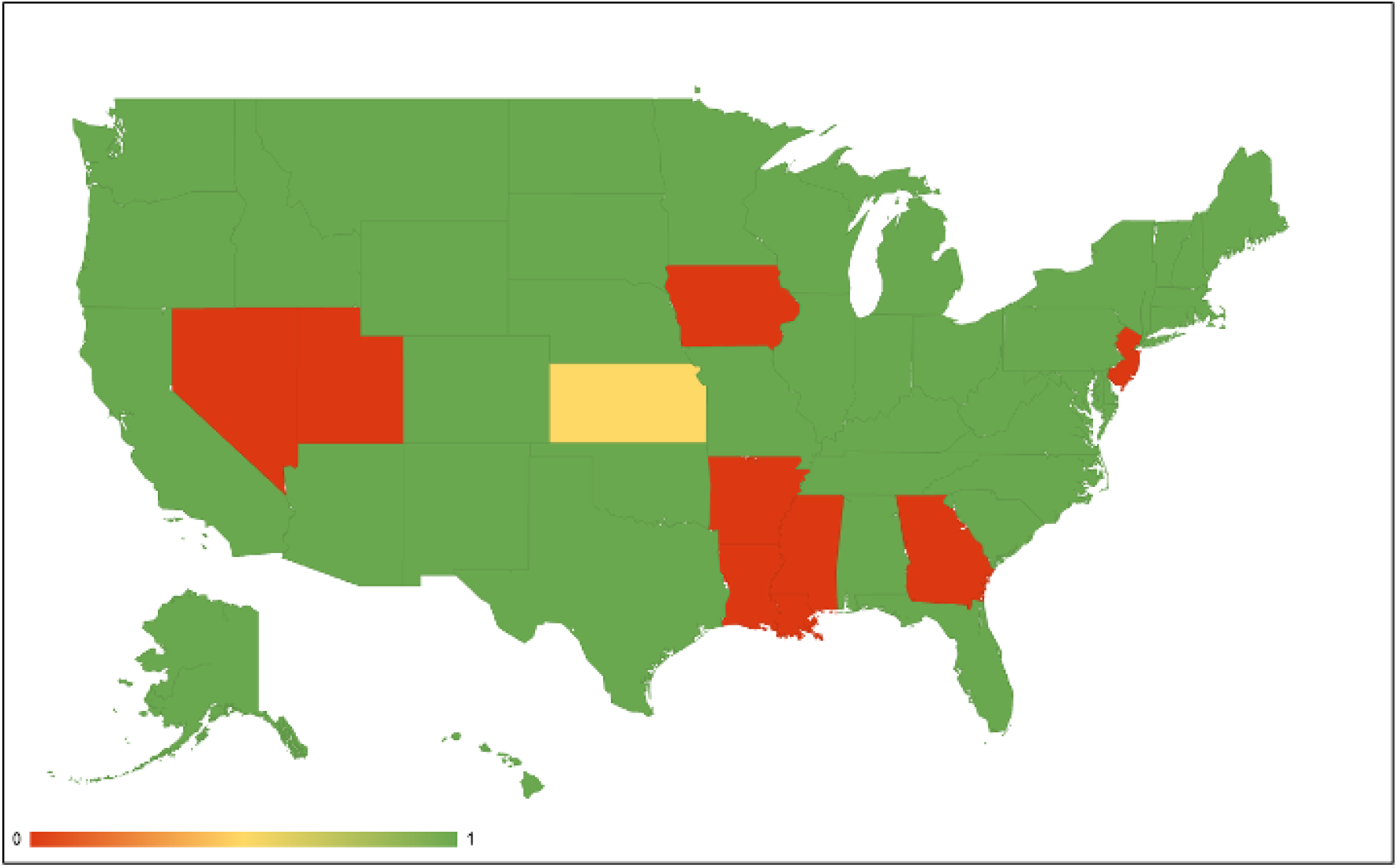
State Medicaid Coverage for Standard Cardiac Rehabilitation Services for state-by-state Medicaid coverage for CPT 93797 (outpatient CR without ECG monitoring). Green = Covered, Yellow = Partial/Inconclusive, Red = Not Covered

The coverage landscape was more limited for CPT code 93798, which includes continuous ECG monitoring and is particularly important for higher-risk patients or those with more complex cardiovascular conditions, including heart failure. Only 43 states provided coverage for this more comprehensive CR service. The same six states: Arkansas, Georgia, Louisiana, Mississippi, Nevada and Utah, also did not reimburse for CPT 93798, and were joined by Kansas as classified as inconclusive, bringing the total number of states lacking full monitored CR coverage to seven (Figure 2).

**Figure 2:**
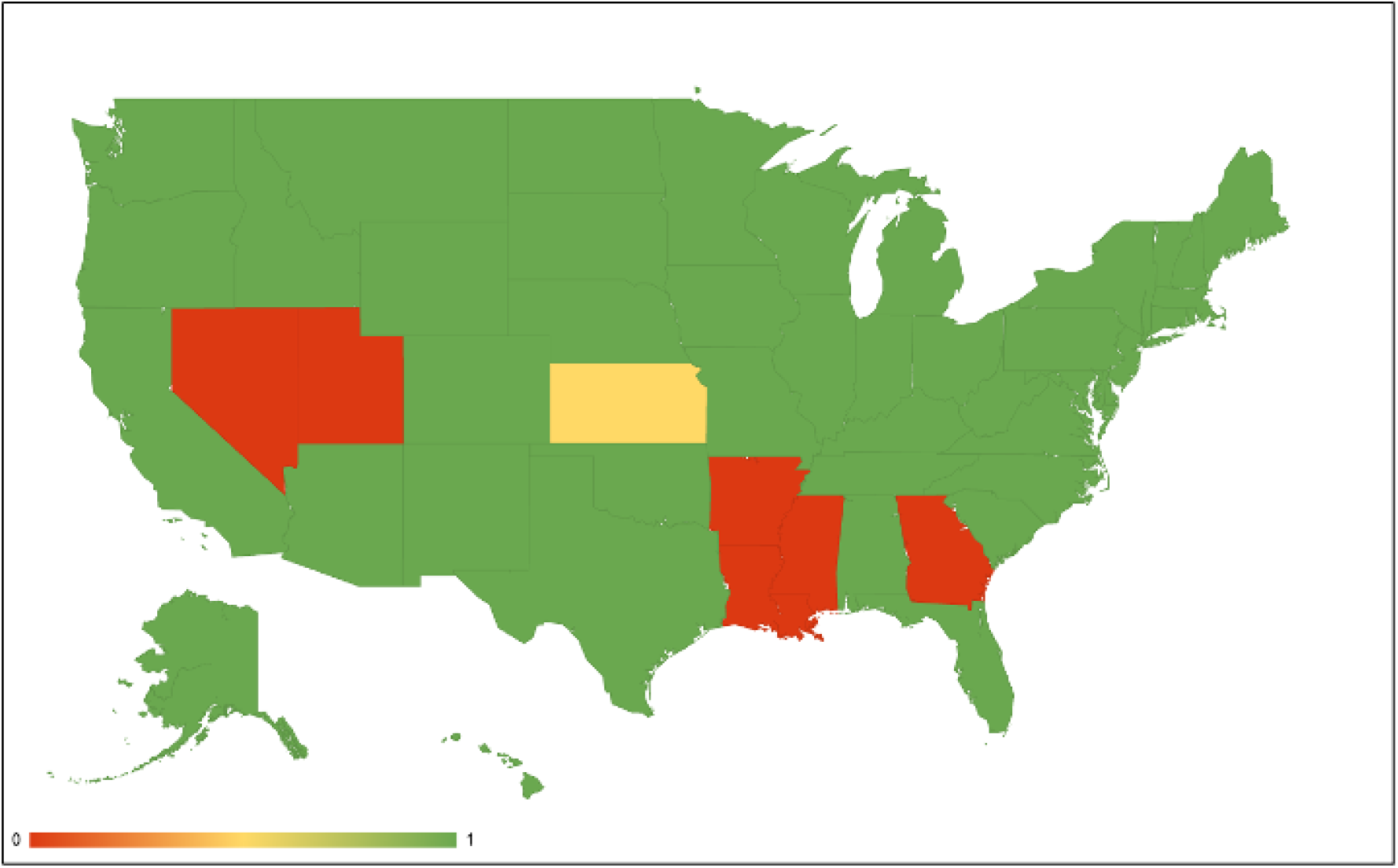
State Medicaid Coverage for Standard Cardiac Rehabilitation Services. Figure 1 shows state-by-state Medicaid coverage for CPT 93798 (outpatient CR with ECG monitoring). Green = Covered, Yellow = Partial/Inconclusive, Red = Not Covered

Figure 3 displays the percentage of Medicaid enrollees by state as of December 2024. States with the highest Medicaid enrollment proportions, exceeding 30% of their total population, include New Mexico, California, and New York. Additional states with enrollment rates between 25% and 30% include Louisiana, Kentucky, West Virginia, and Arkansas. A broader set of states, including Oregon, Arizona, and Mississippi, fall within the 20–25% range. Conversely, states with the lowest Medicaid enrollment percentages under 15% include Utah, Nebraska, North Dakota, and Wyoming, with others such as Idaho and Texas also falling near or below this threshold.

**Figure 3:**
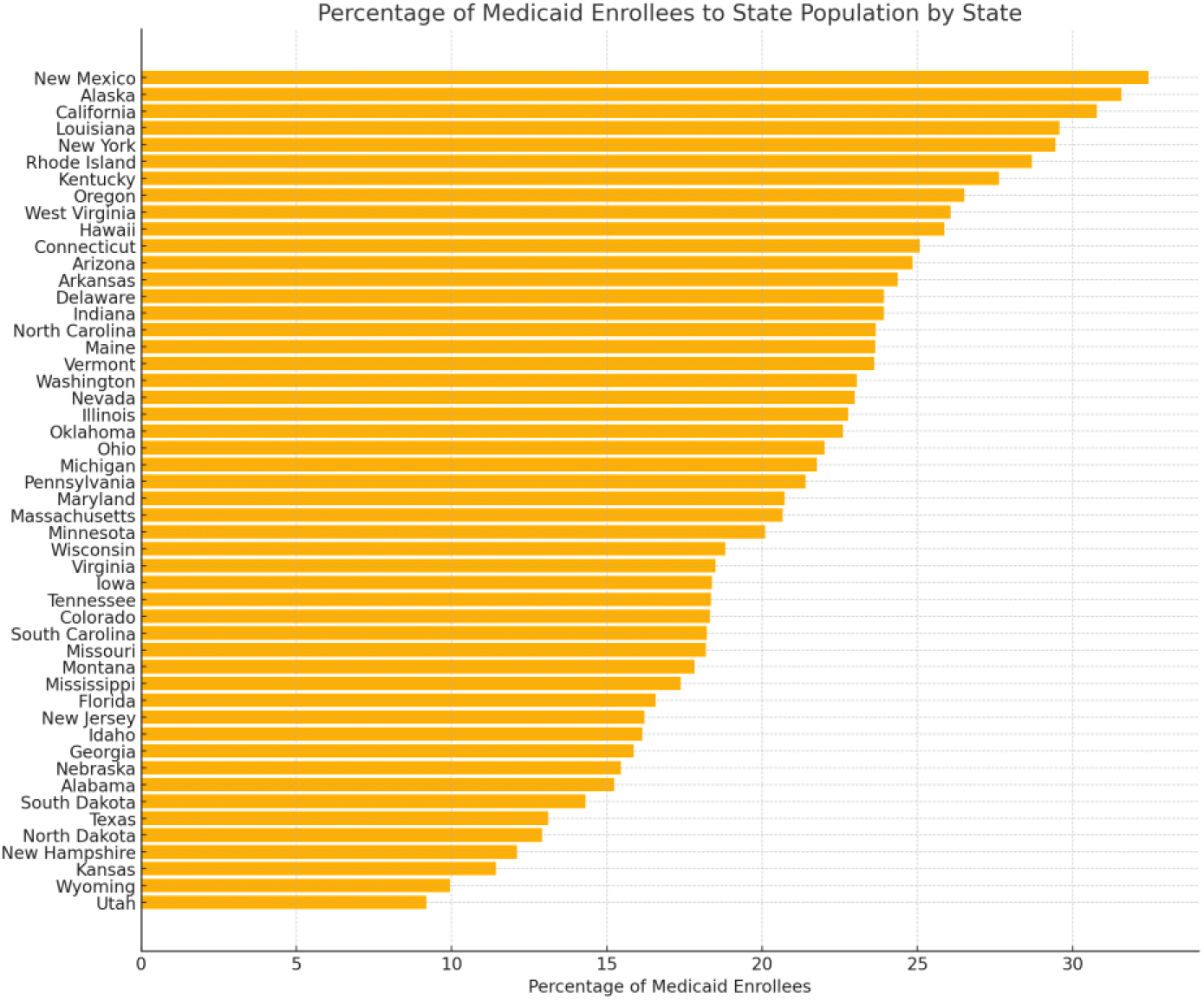
Bar-plot detailing Percentage of US Medicaid Enrollees per State. Data obtained by December 2024 Medicaid & CHIP Enrollment Data Highlights. https://www.medicaid.gov/medicaid/program-information/medicaid-and-chip-enrollment-data/report-highlights

When viewed alongside the cardiac rehabilitation (CR) coverage maps (Figures 2 and 3), the geographic distribution of Medicaid enrollment intersects variably with coverage status. Among high-enrollment states, New York and California provide full Medicaid reimbursement for both CPT 93797 and CPT 93798. In contrast, Louisiana and Arkansas, also with enrollment levels above 25% provide no coverage for either code. Kentucky and West Virginia, also in the high-enrollment category, both offer full CR coverage. Mississippi, with Medicaid enrollment exceeding 20%, does not provide reimbursement for either code.

Of the eight states identified as offering no Medicaid coverage for CPT 93797, Arkansas, Georgia, Iowa, Louisiana, Mississippi, Nevada, New Jersey, and Utah, five (Arkansas, Georgia, Louisiana, Mississippi, and Utah) are also among the seven states not covering CPT 93798. These states are predominantly located in the South (Arkansas, Georgia, Louisiana, Mississippi), the Mountain West (Utah, Nevada), and the Midwest (Iowa). Among these, multiple states have Medicaid enrollment rates exceeding 20%, including Arkansas, Louisiana, and Mississippi.

Geographically, full coverage states are most concentrated in the Northeast (e.g., New York, Massachusetts, Vermont), Upper Midwest (e.g., Minnesota, Michigan), and West (e.g., California, Oregon, Washington). States classified as partial or inconclusive, such as Kansas, tend to fall in the central U.S., while those with no coverage are largely clustered in the South and select Mountain West regions. This distribution reveals a wide spectrum in both CR policy adoption and Medicaid enrollee representation across different U.S. regions.

## Discussion

### Clinical Importance of CR

Cardiac rehabilitation is a clinically validated intervention proven to improve survival and quality of life in patients with HF [3,5]. A Cochrane review encompassing over 6,000 participants found that exercise-based CR reduces all-cause mortality and hospital readmissions while improving health-related quality of life metrics [5]. Moreover, participation in ≥20 sessions yields a significant dose-response relationship, translating into fewer cardiovascular events and enhanced functional status [4,10].

Despite these benefits, CR remains underutilized—particularly among Medicaid beneficiaries, who face a multitude of barriers including limited coverage, transportation issues, and poor referral rates [8,11]. In regions where Medicaid does not cover CR, clinicians are hamstrung in their ability to offer guideline-recommended care, and patients are forced to forego a service that could reduce recurrent hospitalizations and mortality [3,5,6].

### Economic and Policy Implications

From a health economics perspective, failing to provide CR to Medicaid beneficiaries is a shortsighted policy. For every $1 invested in CR, an estimated $4 return is generated through reduced healthcare utilization, particularly by avoiding costly HF rehospitalizations [2,10,16]. States that expanded Medicaid under the ACA have demonstrated significant reductions in cardiovascular mortality, suggesting the broader positive impact of inclusive healthcare policy [9]. From our data, a clear pattern linking Medicaid expansion status under the Affordable Care Act (ACA) with the likelihood of offering full CR coverage was evident. States that had expanded Medicaid eligibility were significantly more likely to provide reimbursement for both CR codes, suggesting a more proactive approach to preventive cardiovascular care in these jurisdictions. These states, concentrated in the Northeastern and Western U.S., have historically demonstrated stronger public health infrastructures and greater investments in chronic disease management. In contrast, non-expansion states—disproportionately located in the South and parts of the Midwest—were overrepresented among those lacking CR coverage, even though these regions bear some of the highest rates of cardiovascular disease, heart failure hospitalizations, and mortality nationwide.

### Health Equity Considerations

Medicaid CR coverage gaps are not only clinical failures but also equity failures compounded disproportionately along socioeconomic, geographic, and racial margins. Disproportionate disease burden in rural, Black, and low-income communities aligns almost exactly with the map of states that exclude CR coverage. For example, Black Americans, who face earlier onset of HF and higher rates of cardiovascular death are less likely to access CR [11,13,14]. Concurrently, patients in rural or impoverished areas may lack transportation to reach a CR facility, cannot afford insurance co-pays, or may work hourly jobs without the flexibility to attend multiple sessions per week [12,15,17]. These social determinants of health (SDOH) act as force multipliers of inequity in regions already burdened by policy exclusion. Equitable cardiovascular care demands not only inclusive insurance policy but proactive elimination of access barriers. Remote and home-based CR programs have shown success in improving participation rates and could be a scalable solution in underserved regions [15,18]. However, this analysis has limitations, as it relies on publicly available policy data and direct agency communication, which may be subject to incomplete or evolving documentation.

### Recommendations

We propose the following actionable steps to address the coverage gap and promote cardiac equity:

1. **Standardize Medicaid coverage for CR (CPT 93797 and 93798) federally** across all states, removing regional variation.
2. **Support expansion of tele-CR programs** to improve rural and transportation-limited access.
3. **Incorporate CR into Medicaid value-based care incentives**, encouraging health systems to emphasize long-term cardiovascular outcomes.
4. **Educate providers on CR referrals and eligibility**, especially in minority-serving and rural institutions.
5. **Encourage CMS to provide direct federal guidance** on CR coverage inclusion for all Medicaid programs.

## Conclusion

Heart failure continues to exact a devastating toll, particularly in underserved and underinsured populations. Cardiac rehabilitation is an underutilized yet essential tool that can reverse this trend—if access is made equitable. By expanding Medicaid coverage to include CR services in every state, we can reduce hospitalizations, improve survival, and move toward a fairer healthcare system.

## Data Availability

The data underlying this study were obtained from publicly available state Medicaid fee schedules, provider manuals, and official state websites. Additional information was obtained through direct communication with state Medicaid agencies. All data used in this analysis are available from the corresponding author upon reasonable request.

## References

1. Heart Failure Society of America. Heart Failure Statistics. https://hfsa.org/patient-hub/heart-failure-facts

2. Kazi DS, et al. Forecasting the economic burden of cardiovascular disease and stroke in the United States through 2050. Circulation. 2024.

3. Bozkurt B, et al. Cardiac rehabilitation for patients with heart failure: JACC expert panel. J Am Coll Cardiol. 2021;77(11):1454–1469.

4. Medina-Inojosa JR, et al. Dose of cardiac rehabilitation to reduce mortality and morbidity: A population-based study. J Am Heart Assoc. 2021;10(20):e021356.

5. Taylor RS, et al. Exercise-based rehabilitation for heart failure. Cochrane Database Syst Rev. 2019;4(4):CD003331.

6. Centers for Medicare & Medicaid Services. Coverage Decision Memo for Cardiac Rehabilitation Programs for Chronic Heart Failure (CAG-00437N). 2014.

7. Osorio A, et al. Medicaid’s Coverage Role in Small Towns and Rural Areas. Georgetown University Center for Children and Families; 2023.

8. Ades PA, et al. Cardiac rehabilitation participation and medical insurance: A review. J Cardiopulm Rehabil Prev. 2017;37(3):125–134.

9. Khatana SAM, et al. Association of Medicaid expansion with cardiovascular mortality. JAMA Cardiol. 2019;4(7):671–679.

10. Thomas RJ, et al. AACVPR/ACCF/AHA 2010 update: performance measures on cardiac rehabilitation. J Am Coll Cardiol. 2010;56(14):1159–1167.

11. Prince DZ, et al. Racial disparities in cardiac rehabilitation initiation and the effect on survival. PM R. 2014;6:486–492.

12. Leung YW, et al. Geographic issues in cardiac rehabilitation utilization: A narrative review. Health Place. 2010;16:1196–1205.

13. Farmer MM, et al. Are racial disparities in health conditional on socioeconomic status? Soc Sci Med. 2005;60:191–204.

14. Karlamangla AS, et al. Socioeconomic and ethnic disparities in cardiovascular risk. Ann Epidemiol. 2010;20:617–628.

15. LaRosa AR, et al. Association of household income and adverse outcomes in atrial fibrillation. Heart. 2020;106:1679–1685.

16. Sérvio TC, et al. Cost and effectiveness assessment of cardiac rehabilitation: a systematic review. Health Econ Rev. 2020;10(1):1–12.

17. Bachmann JM, et al. Factors associated with underutilization of cardiac rehabilitation. Am J Cardiol. 2021;147:82–89.

18. Thomas RJ, et al. Home-based cardiac rehabilitation: a scientific statement. Circulation. 2019;140(1):e69–e89.

